# Do Health Policies Reduce Maternal and Child Mortality in Ghana? Evidence from a Difference-in-Differences Analysis

**DOI:** 10.64898/2026.10.22.26366200

**Authors:** Mawuena Tome, Gideon Ayettey

## Abstract

Ghana introduced a sequence of health policies between 1999 and 2019 to accelerate reductions in maternal and child mortality, including the Community-Based Health Planning and Services initiative, the National Health Insurance Scheme, a Free Maternal Care policy, the Ghana Essential Health Interventions Program, and the National Newborn Health Strategy. Prior evaluations have relied on descriptive trend analysis and cross-sectional comparisons, without constructing a credible counterfactual. This paper applies a staggered 2×2 Difference-in-Difference approach to estimate the causal effect of each policy on annual mortality outcomes over the period 1990 to 2023, comparing Ghana’s trajectory against the global average and the lower-middle-income country group as comparators. Parallel trends are assessed through event-study plots of annual treatment effects. For child mortality, GEHIP (2012) reduced mortality by an estimated 9.5 percent relative to the global trend, and NNHS (2019) by 16.4 percent, though mild pre-existing drift in the NNHS window means the latter is an upper bound. For maternal mortality, GEHIP again produces the most credible estimate at 7.2 percent below the global comparator, supported by a near-flat pre-period ATT series. The Free Maternal Care policy (2008) produced no detectable improvement in maternal mortality relative to either comparator, consistent with evidence that access reforms require adequate facility capacity to translate increased attendance into reduced mortality. CHPS (1999) generates a large apparent maternal mortality effect that is invalidated by a strong pre-existing downward trend in the event-study window. Taken together, the results indicate that supply-side and quality-focused interventions have driven Ghana’s most credible mortality improvements, while financial access reforms have had their strongest effects on child rather than maternal survival.

## Introduction

Ghana occupies a distinctive position in sub-Saharan Africa’s development story. It achieved lower-middle-income status in 2010, sustained a long period of democratic governance, and has been widely cited as a country with functional public institutions [1,2]. In health, these conditions generated real momentum. The country cut its child mortality (defined as death in children under five years [3]) rate by more than half between 1990 and 2015, from around 128 to 60 deaths per 1,000 live births. This is largely achieved through targeted public investment rather than income growth alone [4]. That progress, however, has stalled when it comes to maternal and child mortality outcomes. Recorded maternal deaths have risen sharply in recent years: 819 women died during or shortly after childbirth in 2024, and 504 deaths were recorded in just the first six months of 2025, placing Ghana far from the Sustainable Development Goal targets by 2030 [5]. As of 2025, Ghana’s maternal mortality ratio is estimated at 263 deaths per 100,000 live births, and neonatal mortality stands at 22.8 per 1,000 live births [6]. Projections built on trend data suggest the country’s child mortality rate will reach approximately 30.5 per 1,000 live births by 2030, falling short of the SDG 3 benchmark of 25 or fewer per 1,000 live births [7].

The gap between what has been achieved and what remains to be done in maternal and child health is substantial, and it has persisted despite two decades of active policy intervention.

The Ghanaian government’s response to maternal and child mortality has been extensive. From the late 1990s through the 2010s, the country introduced one major health reform after another. The Community-Based Health Planning and Services initiative went live in 1999, the National Health Insurance Scheme was legislated in 2003 and fully operational by 2004, a Free Maternal Care policy was layered onto the insurance scheme in 2008, a National Ambulance Service was established in 2004, quality improvement protocols for emergency obstetric care were introduced at regional hospitals starting in 2007, the Ghana Essential Health Interventions Program began operating in the Upper West Region around 2012, and a National Newborn Health Strategy was launched in 2019 [4,8–11]. Each of these programs was designed to address a specific bottleneck in the pathway from pregnancy to safe delivery and healthy newborn.

While these successive health policies have had significant impact on the Ghanaian health system, their impact on maternal and child mortality are under explored. Most studies evaluate a single program, compare outcomes before and after the policy introduction without constructing a credible counterfactual, or describe trends without attempting to isolate a causal effect [4,8].

The difficulty is real: policies overlap in time and concurrent changes in education, income, and fertility all move outcomes in ways that are hard to disentangle from the policies themselves.

Without a rigorous identification strategy, it is not possible to say with confidence whether any observed decline in mortality after a policy change is due to the policy or to some other trend that would have continued regardless. This paper addresses that limitation directly. Using a Difference-in-Difference approach that exploits variation in the timing and implementation of key health policies in Ghana, we estimate the causal effect of these interventions on maternal and infant mortality. To our knowledge, no existing study applies this identification strategy to the joint evaluation of Ghana’s maternal and child health policy portfolio.

The leading causes of maternal death in Ghana are obstetric haemorrhage (22.8 percent of maternal deaths), abortion complications (13.7 percent), infectious diseases including sepsis (13.9 and 6.9 percent respectively), hypertensive disorders (8.9 percent), and obstructed labour (4.5 percent) [12]. Most are clinically manageable when the health system can deliver timely skilled interventions: haemorrhage with uterotonics and surgical capacity, eclampsia with magnesium sulfate, sepsis with antibiotics, and obstructed labour with caesarean section [10,13].

Women in rural areas and those without formal education face systematically higher risks, meaning geographic access to quality care is the binding constraint that the policies under study were designed to address [12,14].

Child mortality accounts for a growing share of children deaths in Ghana as the decline in child mortality has been slower than for older children [4]. The leading causes are preterm birth complications (49.6 percent), birth asphyxia (21.7 percent), neonatal infections (14.6 percent), and congenital anomalies (8.2 percent) [15]. Care-seeking delays, inadequate antenatal attendance, and the absence of immediate newborn care compound these clinical risks, particularly in rural settings [11,14]. Ghana is projected to approach but not reach the SDG 3 child mortality target of 25 deaths per 1,000 live births by 2030 without additional intervention [7].

Ghana’s response to maternal and child mortality unfolded in successive waves, with Table 1 summarizing the full portfolio. CHPS (1999) placed community health officers in rural villages to close the spatial gap between women and skilled care; national evaluation found that CHPS paired with a nearby facility increased skilled birth attendance from 36.4 to 48.5 percent, though CHPS alone, without facility access, produced no significant effect [8]. The NHIS (2003), Ghana’s most comprehensive financial reform, reduced out-of-pocket costs for prenatal care, delivery, malaria, and other essential services, and has been associated with higher antenatal attendance and institutional delivery [9,16]. The Free Maternal Care policy (2008) extended that logic by eliminating residual user fees specifically for antenatal care, delivery, and postnatal visits, targeting women for whom even small payments remained prohibitive [9]. GEHIP (2012) took a different approach, bundling supply-side facility strengthening and service integration at the district level in the Upper West Region, where mortality burdens were highest and health system capacity most constrained [14]. Supply-side factors are the structural capacity of a health system, such as the availability of facilities, staff, supplies, and functioning institutions to deliver care [17]. The National Newborn Health Strategy (2019) was designed in response to evidence that neonatal mortality had been declining more slowly than other child mortality indicators, and directly targets preterm birth complications, birth asphyxia, neonatal sepsis, and congenital anomalies [4,11].

**Table 1:** Ghana Health Policies Targeting Maternal and Child Mortality, 1999-2019.

| Policy | Year | Primary Causes Addressed |
| --- | --- | --- |
| Community-Based Health Planning and Services (CHPS) | 1999 | Haemorrhage, sepsis, obstructed labour, malaria, neonatal care (via improved access to skilled birth care) |
| National Health Insurance Scheme (NHIS) | 2003/2004 | Malaria, hypertension, diabetes, prenatal complications, haemorrhage, normal and complicated delivery |
| User Fees Exemption for Delivery (UFEDC) | 2004 | All causes of maternal death at delivery (financial barrier removal) |
| Focused Antenatal Care (FANC) | 2004 | Hypertension, anaemia, infections, pregnancy complications (early detection) |
| National Ambulance Service (NAS) | 2004 | Haemorrhage, obstructed labour, eclampsia (emergency transport delays) |
| Free Maternal Care Policy | 2008 | All direct maternal causes: haemorrhage, eclampsia, sepsis, obstructed labour, abortion complications |
| Emergency Obstetric and Newborn Care (EmONC) Quality Improvement | 2007 | Obstetric haemorrhage, hypertensive disorders, sepsis, obstructed labour, birth asphyxia |
| Ghana Essential Health Interventions Program (GEHIP) | 2012 | Haemorrhage, sepsis, obstructed labour, malaria, child illness (Upper West Region) |
| MDG 5 Accountability Framework | 2010 | All maternal mortality causes (accountability and reporting structure) |
| National Newborn Health Strategy (NNHS) | 2019 | Prematurity, birth asphyxia, neonatal sepsis, congenital anomalies |

Note: Policy years and descriptions compiled from Kayode et al. (2016), Johnson et al. (2015), Brugiavini and Pace (2016), Srofenyoh et al. (2016), and Adongo and Ganle (2023).

The evidence that each program improved utilization or intermediate outcomes is consistent with the existing literature. For example, CHPS increased skilled birth attendance when paired with facilities [8]. NHIS increased antenatal attendance and institutional delivery [9,16]. Quality improvement at Ridge Regional Hospital reduced maternal case fatality rates from obstetric haemorrhage by more than 90 percent between 2007 and 2011, documenting one of the clearest facility-level causal chains in the Ghanaian literature [10]. The policies described above represent a substantial and sustained national investment. Quality improvement at the facility level measurably reduced case fatality rates [10]. For example, neonatal skin-to-skin care is associated with dramatically lower odds of death [11].

What the literature has not established is the causal effect of these policies on population-level mortality. Kayode et al. [4] note that mortality changes in Ghana during this period cannot be attributed directly to specific policies given overlapping implementation and insufficient identification strategies.

This paper makes three contributions. Methodologically, we apply a staggered 2×2 Difference-in-Difference framework that evaluates five policies in sequence, each within its own non-overlapping pre-and post-implementation window, avoiding the contaminated weighted averages produced by pooled two-way fixed effects estimators in staggered adoption settings [18–20]. Event-study plots for each window provide a visual basis for assessing whether parallel trends hold before each intervention. In terms of scope, no existing study jointly evaluates the full sequence of major Ghanaian health interventions from CHPS (1999) through NNHS (2019) on both maternal and child mortality within the same framework, which allows us to ask whether financial access reforms and supply-side quality programs have systematically different effects on these two outcomes. Substantively, the results show that the most credible mortality improvements are associated with supply-side programs: GEHIP (2012) generates the most defensible estimates for both outcomes, while the Free Maternal Care policy (2008) produced no detectable change in maternal mortality, with direct implications for how policymakers in comparable settings should sequence access and quality investments.

## Methodology

This paper uses two annual time series for Ghana drawn from the World Bank World Development Indicators database, last updated April 8, 2026. Both series cover 1990 through 2023, providing a 34-year window that spans a pre-policy baseline of nine years before the first major intervention in 1999 and the full period of subsequent reforms through 2023. The child mortality estimates extend further back in history, but the analysis uses 1990 as the common start year so that both outcomes are evaluated over the same observation window and compared against the same set of global and income-group benchmarks.

The first outcome is Ghana’s child mortality rate, i.e. the under-five mortality rate (U5MR), measured as deaths per 1,000 live births (World Bank indicator SH.DYN.MORT), sourced from the UN Inter-agency Group for Child Mortality Estimation (IGME), which harmonizes vital registration, household surveys, and demographic surveillance data into a consistent annual series. Ghana’s U5MR stood at 127.6 in 1990, declined to 103.9 by 1999 when CHPS launched, and reached 35.9 by 2023, highlighting a 72 percent reduction over the full window. The nine years from 1990 to 1998 serve as the pre-policy baseline.

The second outcome is Ghana’s maternal mortality ratio (MMR), measured as deaths per 100,000 live births (World Bank indicator SH.STA.MMRT), sourced from the WHO-led *Trends in Maternal Mortality* series. Because maternal deaths are rare relative to the population at risk, the series uses a regression model drawing on the proportion of maternal deaths among non-AIDS deaths in women aged 15–49, the total fertility rate, skilled birth attendance, and GDP per capita. Ghana’s MMR was 943 in 1990, fell to 630 by 1999, and reached 234 by 2023. Both series cover 1990–2023, with nine pre-CHPS baseline observations.

Table 2 reports full descriptive statistics for both outcomes across all four series. Three features are worth highlighting. First, Ghana’s child mortality rate mean (79.3) tracks the lower-middle-income average (80.3) closely, confirming that Ghana has broadly kept pace with its income-group peers on child survival, though it still lags the global average of 62.4 due to the weight of high-income countries in that aggregate. For maternal mortality, Ghana’s mean of 429.6 exceeds both the global (287.9) and lower-middle-income (375.9) averages, indicating weaker relative performance on that outcome. Second, ADF and PP tests reject the unit root for Ghana on both outcomes while the KPSS simultaneously rejects stationarity, pointing to trend-stationarity: both series follow deterministic downward trends rather than random walks. Third, Ljung-Box Q(20) statistics are significant at the 1 percent level across all series.

**Table 2:** Descriptive Statistics of the Variables.

| Statistic | Child Mortality Rate |  |  |  | Maternal Mortality Ratio |  |  |  |
| --- | --- | --- | --- | --- | --- | --- | --- | --- |
|  | Ghana | Global Average | SSF Average | Lower Middle Income | Ghana | Global Average | SSF Average | Lower Middle Income |
| Mean | 79.312 | 62.368 | 121.450 | 80.321 | 429.618 | 287.882 | 694.265 | 375.912 |
| Variance | 837.475 | 394.547 | 1474.493 | 731.309 | 24590.243 | 4383.804 | 25528.382 | 10425.659 |
| Std. Dev. | 28.939 | 19.863 | 38.399 | 27.043 | 156.813 | 66.210 | 159.776 | 102.106 |
| Skewness | 0.115<br>(0.753) | 0.272<br>(0.459) | 0.272<br>(0.458) | 0.305<br>(0.408) | 0.785**<br>(0.045) | 0.321<br>(0.384) | 0.475<br>(0.205) | 0.442<br>(0.236) |
| Kurtosis | 1.639***<br>(0.001) | 1.533***<br>(0.000) | 1.496***<br>(0.000) | 1.704***<br>(0.002) | 2.491<br>(0.697) | 1.683***<br>(0.002) | 2.261<br>(0.353) | 1.922**<br>(0.043) |
| ADF | -6.118***<br>(0.000) | -1.991<br>(0.290) | -1.010<br>(0.750) | -2.935**<br>(0.041) | -5.368***<br>(0.000) | -1.762<br>(0.400) | -1.423<br>(0.571) | -3.527***<br>(0.007) |
| PP | -3.510***<br>(0.008) | -2.904**<br>(0.045) | -2.131<br>(0.232) | -2.975**<br>(0.037) | -2.848*<br>(0.052) | -2.216<br>(0.201) | -2.211<br>(0.202) | -2.382<br>(0.147) |
| KPSS | 0.785**<br>(0.010) | 0.763**<br>(0.010) | 0.760**<br>(0.010) | 0.777**<br>(0.010) | 0.742**<br>(0.010) | 0.765**<br>(0.010) | 0.761**<br>(0.010) | 0.773**<br>(0.010) |
| JB | 2.700<br>(0.259) | 3.468<br>(0.177) | 3.626<br>(0.163) | 2.905<br>(0.234) | 3.861<br>(0.145) | 3.040<br>(0.219) | 2.049<br>(0.359) | 2.755<br>(0.252) |
|  | Ghana | Global Average | SSF Average | Lower Middle Income | Ghana | Global Average | SSF Average | Lower Middle Income |
| Q(20) | 190.365***<br>(0.000) | 210.953***<br>(0.000) | 213.052***<br>(0.000) | 188.566***<br>(0.000) | 128.829***<br>(0.000) | 191.300***<br>(0.000) | 124.826***<br>(0.000) | 169.279***<br>(0.000) |
| Q<sup>2</sup>(20) | 184.475***<br>(0.000) | 199.945***<br>(0.000) | 204.134***<br>(0.000) | 176.197***<br>(0.000) | 113.142***<br>(0.000) | 184.764***<br>(0.000) | 120.753***<br>(0.000) | 158.619***<br>(0.000) |

Note: Both panels cover 1990-2023. Child mortality is expressed as deaths per 1,000 live births; maternal mortality ratio as deaths per 100,000 live births. Std. Dev. denotes standard deviation. ADF = Augmented Dickey-Fuller test; PP = Phillips-Perron test; KPSS = Kwiatkowski-Phillips-Schmidt-Shin test; JB = Jarque-Bera normality test. Q(20) and Q<sup>2</sup>(20) denote Ljung-Box statistics for the series and squared series up to 20 lags. KPSS p-values are bounded at 0.010 and 0.100 by the tabulated critical values. SSF = Sub-Saharan Africa. Data source: World Bank WDI (SH.DYN.MORT and SH.STA.MMRT), last updated April 8, 2026. \*, \*\*, \*\*\* indicate significance at the 10%, 5%, and 1% levels. p-values in parentheses.

Figure 1 shows all four child mortality rate series on a consistent downward path since 1990. Ghana entered the period at 127.6 deaths per 1,000 live births, nearly identical to the lower-middle-income average of 127.9, and both declined in close proximity. By 2023 Ghana reached 37.2, marginally below the global average of 38.3 and well below the lower-middle-income group at 44.9. The steepest declines coincide with the period of intensified policy activity from CHPS (1999) through Free MC (2008), after which the pace of decline slowed across all series. This is consistent with diminishing returns as the most preventable deaths are addressed first. Ghana has not yet reached the SDG 3 target of 25 deaths per 1,000 live births.

**Figure 1:**
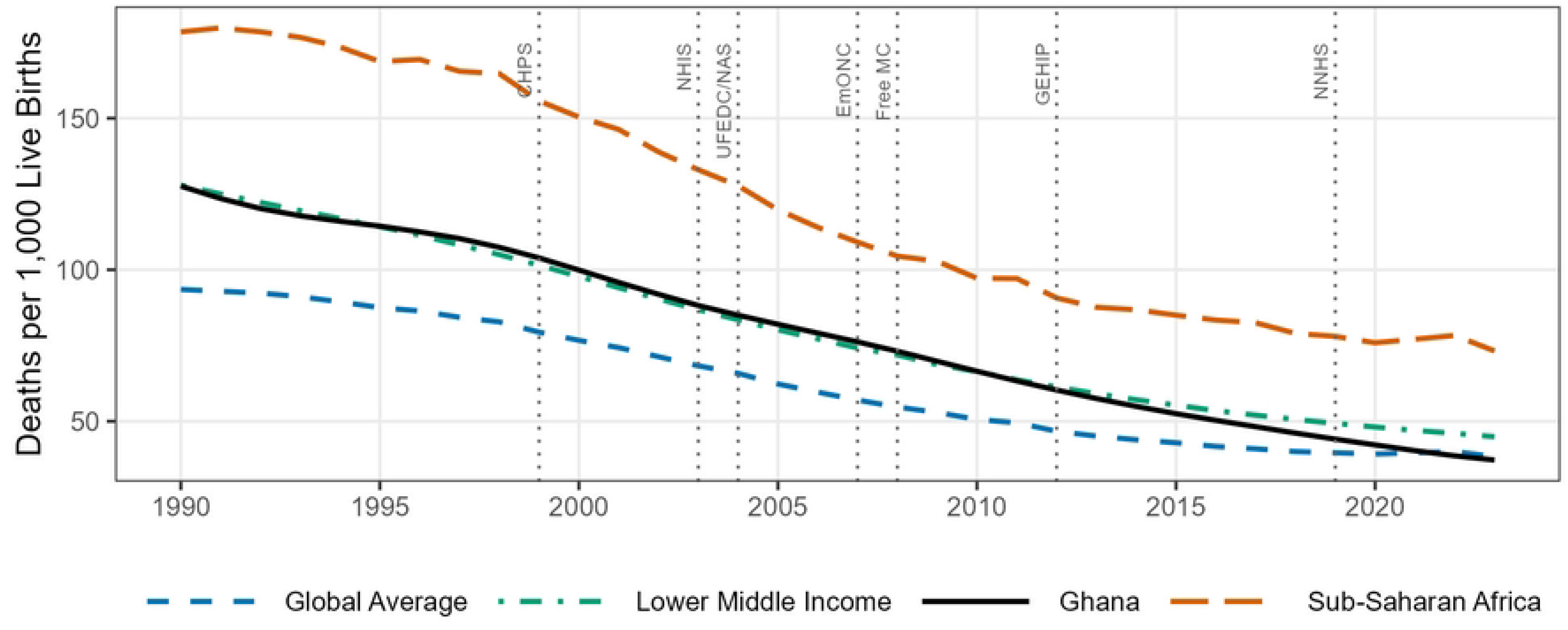
Child Mortality Rate in Ghana and Comparator Groups, 1990-2023

Figure 2 tells a different story for maternal mortality. Ghana started 1990 at 769 deaths per 100,000 live births, well above the global average of 391 and the lower-middle-income average of 572. By 2023, Ghana’s ratio stood at 234, converging to the lower-middle-income average of 235 after more than three decades of decline, and still above the global average of 197. A visible uptick around 2021 is consistent with COVID-19 disruption to maternal health services, with recovery by 2023. The more gradual pace of decline since 2008 relative to the 1990s suggests persistent gaps in obstetric care quality that financial access reforms alone have not been enough to close.

**Figure 2:**
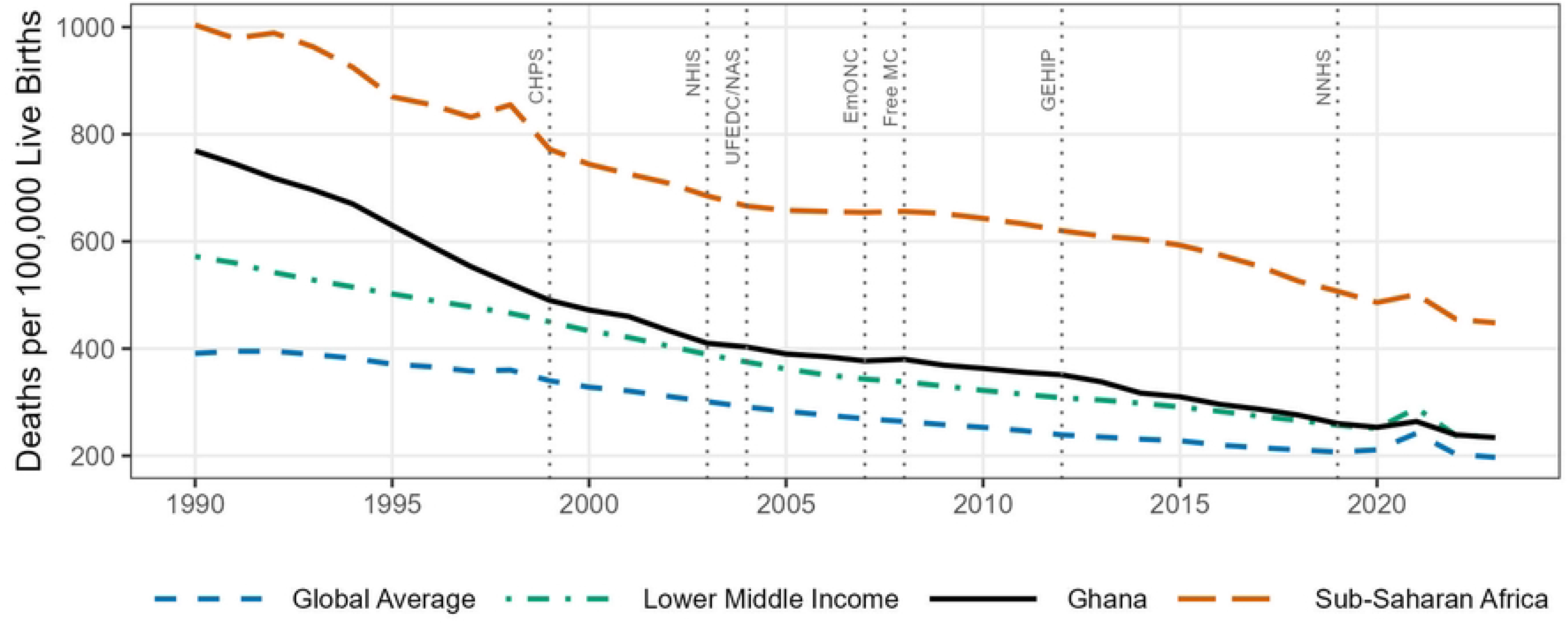
Maternal Mortality Ratio in Ghana and Comparator Groups, 1990-2023

The Difference-in-Difference approach below relies on three comparator groups, the global average (World Bank code WLD), the Sub-Saharan African average (SSF), and the lower-middle-income country average (LMC), each of which is a World Bank aggregate published across dozens to hundreds of constituent countries. Because Ghana is itself a member of both the lower-middle-income income group and the Sub-Saharan African region, its own mortality figures enter the LMC and SSF aggregates, and contribute marginally to the WLD aggregate as well. This creates a mild source of circularity in principle: Ghana’s change in mortality is being measured against a benchmark that partly contains Ghana’s own change. To assess whether this materially affects the results, we recomputed each comparator group directly from the underlying country-level WDI series, using the World Bank’s Region and IncomeGroup classifications to identify the 50 countries in the LMC group, the 47 countries in the SSF group, and the 215 countries in the WLD group, and compared the resulting averages with and without Ghana included. Across both outcomes and all three comparators, dropping Ghana shifts the full-period mean by less than one percent in every case, and the 2023 cross-sectional values move by a similarly negligible margin. This is unsurprising given the scale involved: Ghana’s population of roughly 33 million is a small fraction of the LMC group’s total, which includes India, Indonesia, Pakistan, and Vietnam among its members, so even a single country with a meaningfully different mortality trajectory cannot move an average built from dozens to hundreds of countries by more than a trivial amount. We therefore conclude that the inclusion of Ghana in its own comparator groups does not introduce any economically significant bias into the DiD estimates, and none of the paper’s substantive conclusions would change under a leave-Ghana-out construction of the comparators.

Ghana’s mortality rates fell over the entire 1990 to 2023 period, and many things changed alongside the policies: income per capita grew, educational attainment improved, fertility declined, and global child health interventions expanded [21]. A simple before-and-after comparison of Ghana’s mortality trends would conflate the effects of specific policies with these background changes. To separate what the policies contributed from what would have happened anyway, this paper uses a Difference-in-Difference framework that compares Ghana’s change in mortality around each policy event to the change in mortality observed in three comparison groups over the same interval.

The comparison groups are the global average (World Bank code WLD), the Sub-Saharan African average (SSF), and the lower-middle-income country average (LMC). None of these groups received the specific Ghanaian health policies, so their mortality trends over any given policy window provide an estimate of the counterfactual trend, that is, how Ghana’s mortality would have moved had the policy not been introduced. Ghana’s actual change in mortality over the same window, minus the comparator’s change, gives the policy’s estimated causal effect.

The approach follows the forward-engineering logic of Baker et al. [20] and Callaway and Sant’Anna [19], which organize complex multi-period DiD studies as collections of simple two-by-two building blocks rather than as a single pooled two-way fixed effects (TWFE) regression. A pooled TWFE model with all five policy dummies stacked into one equation would produce a contaminated weighted average of treatment effects, because earlier policy estimates would use later post-treatment observations as their comparison periods. This problem with conventional TWFE in settings with staggered adoption and heterogeneous treatment effects is documented by Goodman-Bacon [18], De Chaisemartin and D’Haultfœuille [22], Sun and Abraham [23], and Borusyak et al. [24], whose collective findings motivate the building block structure used here.

The building block approach avoids contamination by estimating each policy’s effect separately within a clean, non-overlapping pre-and post-treatment window. The five policies studied here, their implementation years, and their designated estimation windows are described in Table 3.

**Table 3:** Staggered 2×2 DiD Estimation Windows by Policy.

| Policy | Year | Pre-Treatment Window | Post-Treatment Window |
| --- | --- | --- | --- |
| Community-Based Health Planning and Services (CHPS) | 1999 | 1990-1998 | 1999-2002 |
| National Health Insurance Scheme (NHIS) | 2003 | 1999-2002 | 2003-2007 |
| Free Maternal Care Policy (Free MC) | 2008 | 2003-2007 | 2008-2011 |
| Ghana Essential Health Interventions Program (GEHIP) | 2012 | 2008-2011 | 2012-2018 |
| National Newborn Health Strategy (NNHS) | 2019 | 2012-2018 | 2019-2023 |

Note: The pre-treatment window for each policy ends the year before implementation. The post-treatment window ends the year before the next policy in the sequence. NNHS post-treatment window runs to the last year of available data (2023). Windows are non-overlapping by construction to prevent contamination across policy estimates.

The policy window boundaries reflect two design choices. First, each pre-treatment window ends the year immediately before a policy takes effect, to avoid including any anticipation effects in the baseline. Second, each post-treatment window ends the year before the next policy is introduced, so the estimated effect of one policy is not contaminated by the contemporaneous arrival of the next. CHPS (1999) uses the full period from 1990 to 1998 as its pre-treatment baseline because this is the earliest year for which comparable data exist across all four series.

NNHS (2019) uses all available years through 2023 as its post-treatment window, since no subsequent policy displaces it. The policies were selected on the basis of documented national coverage and meaningful year spacing, which ensures that each window contains enough observations to characterize the pre-treatment trend and measure the post-treatment change.

### Estimation Framework

For each policy *g* ∈ {1999,2003,2008,2012,2019} and each comparator *c* ∈ {WLD,SSF,LMC}, the DiD estimator takes the following form:

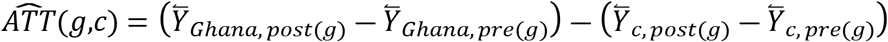

where *Ȳ_G_*_ℎ*ana*,_ *_post_*_(*g*)_ is the mean of the outcome variable for Ghana over the post-treatment window defined for policy *g*, *Ȳ_G_*_ℎ*ana*,_ *_pre_*_(*g*)_ is the mean of the outcome variable for Ghana over the corresponding pre-treatment window, and *Ȳ_c_*, *_post_*_(*g*)_ and *Ȳ_c_*, *_pre_*_(*g*)_ are the analogous means for comparator group *c* over the same windows. The estimator *AT͡T*(*g*,*c*) is the average treatment effect on the treated for policy *g*, estimated using comparator *c*. It measures how much the outcome changed for Ghana above and beyond the change observed in the comparator, removing the common trend component from the estimate.

The outcome variable *Y* is either the child mortality rate (deaths per 1,000 live births) or the maternal mortality ratio (deaths per 100,000 live births), evaluated separately. The two outcomes are not combined because they respond to different subsets of the policies in Table 3: CHPS, NHIS, and GEHIP operate through channels that affect both child and maternal outcomes, while Free MC and NNHS are more directly targeted at maternal and neonatal pathways respectively. To express the estimated effect in percentage terms, the level estimate is divided by Ghana’s pre-treatment mean for that policy window:

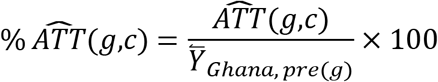

This percentage figure answers the question: by how many percent of its pre-policy baseline did Ghana’s mortality rate change above and beyond the trend in the comparison group? A negative value means Ghana’s mortality fell faster than the comparator over that window, which is interpreted as evidence that the policy contributed to mortality reduction. A positive value would mean Ghana’s mortality fell more slowly than the comparator, suggesting the policy either had no effect or was introduced during a period when Ghana was falling behind.

Each policy is estimated three times, once per comparator, producing a range of point estimates. The Global Average and the Lower-Middle-Income average are treated as the primary comparators because they track income-comparable populations and follow trajectories that are more plausibly parallel to Ghana’s over most of the sample. Sub-Saharan Africa entered 1990 at a much higher mortality level and with a different slope, so the parallel trends assumption is less likely to hold for that series, particularly for pre-treatment windows early in the sample. The SSF estimates are therefore reported as secondary robustness checks rather than as the main identification basis.

### Identification

The identifying assumption behind each *ATT*(*g*,*c*) is parallel trends: in the absence of policy *g*, Ghana’s mortality would have changed by the same amount as comparator *c* over the estimation window [20]. This assumption is stated formally as:

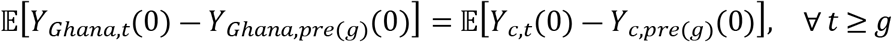

where *Y_G_*_ℎ*ana*,*t*_(0) denotes Ghana’s potential outcome at time *t* in the absence of the policy, and *Y_c_*_,*t*_(0) is the comparator’s outcome in period *t*. Neither of these counterfactual quantities is observed after the policy takes effect, so the assumption cannot be tested directly in post-treatment periods. What can be tested is whether the two series were moving in parallel before the policy. For each estimation window, a pre-trend check compares the slope of Ghana’s outcome trend in the pre-treatment years to the slope of the comparator’s trend over the same period. If the pre-treatment trends are close and not statistically distinguishable, this provides some confidence that the parallel trends assumption is reasonable, though it does not prove it holds post-treatment.

The second identifying assumption is no anticipation: Ghana’s mortality should not have changed in response to a policy before the policy was formally in place. If health workers or households adjusted their behavior in the years immediately before a policy launch, the pre-treatment period would already contain part of the treatment effect, and the estimated *ATT*(*g*,*c*) would be smaller in absolute terms than the true effect. For each policy, the pre-treatment window is therefore defined to end one year before the formal implementation date, and any visible break in Ghana’s pre-treatment trend in the final year of that window is noted as a potential anticipation concern.

Three identification caveats apply. First, policies overlap in their mechanisms even when their estimation windows do not: effects attributed to later policies may partly reflect the continuation of channels opened by earlier ones rather than wholly independent contributions. Second, the comparator groups themselves contain countries implementing their own health reforms during this period; where those reforms coincide with Ghana’s, the comparator trend absorbs some of the policy effect, making the *ATT*(*g*,*c*) estimates conservative lower bounds on true policy impacts. Third, both outcome series are modeled estimates that impose smoothness on the underlying data, which may attenuate the visible breaks around policy years and reduce the informativeness of pre-trend assessments relative to what raw administrative counts would provide.

## Results

Table 4 reports the main DiD estimates. For each policy the table shows the ATT in levels and as a percentage of Ghana’s pre-policy mean, estimated separately against the Global Average and the Lower Middle Income comparator. Negative values indicate that Ghana’s mortality fell faster than the comparator over the relevant window, which is interpreted as a policy-attributable improvement above and beyond the background trend.

**Table 4:** Staggered 2×2 DiD Estimates: Average Treatment Effects on Child Mortality and Maternal Mortality.

| Policy | Child Mortality Rate |  | Maternal Mortality Ratio |  |
| --- | --- | --- | --- | --- |
|  | vs. Global Average | vs. Lower Middle Income | vs. Global Average | vs. Lower Middle Income |
| CHPS (1999) | -5.272 (-4.52%) | +1.992 (+1.71%) | -137.222 (-20.96%) | -101.028 (-15.43%) |
| NHIS (2003) | -2.950 (-3.01%) | -0.205 (-0.21%) | -30.000 (-6.47%) | -7.750 (-1.67%) |
| Free MC (2008) | -3.250 (-3.96%) | -1.220 (-1.49%) | +2.500 (+0.64%) | +11.750 (+2.99%) |
| GEHIP (2012) | -6.450 (-9.46%) | -3.229 (-4.74%) | -26.357 (-7.18%) | -19.179 (-5.23%) |
| NNHS (2019) | -8.660 (-16.39%) | -3.851 (-7.29%) | -47.143 (-15.17%) | -25.571 (-8.23%) |

Note: Each cell reports the ATT in levels followed by the percentage effect relative to Ghana's pre-policy mean in parentheses. A negative value indicates Ghana's mortality fell faster than the comparator over the estimation window. ATT for Child Mortality Rate is in deaths per 1,000 live births; ATT for Maternal Mortality Ratio is in deaths per 100,000 live births. Global Average = World Bank WLD aggregate; Lower Middle Income = World Bank LMC aggregate. Sub-Saharan Africa (SSF) robustness estimates are reported in Table A1.

The results for child mortality are most consistent for the later policies. GEHIP (2012) is associated with a reduction of 6.45 deaths per 1,000 live births relative to the global trend, equivalent to 9.5 percent of Ghana’s pre-GEHIP mean of 68.2, and a reduction of 3.23 deaths relative to lower-middle-income peers, equivalent to 4.7 percent. NNHS (2019) shows the largest percentage effect on child mortality rate in the full table, with a 16.4 percent improvement relative to the Global Average and 7.3 percent relative to both the LMC and the Sub-Saharan African averages (see Table A1). Given that both primary comparators point in the same direction and are of similar magnitude for NNHS adds confidence to the estimate. Free MC (2008) also shows consistent negative ATTs across both primary comparators, at 4.0 percent relative to Global and 1.5 percent relative to LMC, though the effect is smaller and the two comparators do not agree as closely in magnitude. NHIS (2003) shows a modest 3.0 percent improvement relative to the Global Average but almost no difference relative to the LMC group, which is the more appropriate income-group benchmark; this is consistent with NHIS being primarily a financial access reform rather than a supply-side child health intervention. CHPS (1999) shows a negative ATT versus the Global Average but a small positive value versus LMC, meaning that Ghana’s child mortality rate fell at roughly the same pace as lower-middle-income peers over the 1999 to 2002 window, and the apparent improvement relative to the global series reflects a widening gap between rich and poor countries rather than a Ghana-specific gain.

For maternal mortality, the picture is more varied. CHPS (1999) produces the largest level ATT in the full table at ― 137.2 deaths per 100,000 relative to the Global Average, a 21.0 percent reduction, and ― 101.0 relative to LMC, a 15.4 percent reduction. These are large numbers, but the event-study plot in Figure 3 shows a strong downward pre-trend in the annual ATT series throughout the 1990 to 1998 pre-window: the differential gap between Ghana and the global average was narrowing by roughly 12 to 15 deaths per year even before CHPS was introduced. The pre-trend is not flat, which means the parallel trends assumption is unlikely to hold for this window, and the CHPS estimate for MMR should be treated with caution. It likely captures the tail end of a steep pre-existing decline rather than a policy-induced shift. NHIS (2003) shows a moderate 6.5 percent improvement relative to Global and 1.7 percent relative to LMC. The pre-trends for the NHIS window are somewhat better behaved, with the annual ATT moving from + 11.0 in 1999 to ― 16.0 in 2002, a trend that moderates relative to the CHPS window, though it is not fully flat. Free MC (2008) is the only policy that produces a positive ATT against both primary comparators, at + 0.6 percent versus Global and + 3.0 percent versus LMC, meaning that Ghana’s MMR declined more slowly than these groups during the 2008 to 2011 window. The event-study plot in Figure 4 shows that the annual ATT for this window was near zero throughout, with no systematic downward shift after 2008. This is an unexpected finding given that Free MC was designed specifically to remove financial barriers to maternity care. The result is consistent with evidence that access reforms work only when facility quality is sufficient to convert attendance into survival, and it aligns with Srofenyoh et al. [10], who documented that emergency obstetric care quality at the facility level remained a binding constraint through this period. GEHIP (2012) recovers a moderate 7.2 percent improvement relative to Global and 5.2 percent relative to LMC on MMR. The pre-trend for this window is nearly flat (annual ATT moves from + 4.5 to ― 2.5 over the pre-period), which is the best pre-trend behavior of any MMR window and makes this the most credible MMR estimate in the table. NNHS (2019) generates a large 15.2 percent improvement against the Global Average and 8.2 percent against LMC. The event-study path accelerates sharply through 2020 and 2021 before pulling back in 2022 and 2023, which partly reflects the COVID-19 disruption and the subsequent recovery visible in the descriptive data.

**Figure 3:**
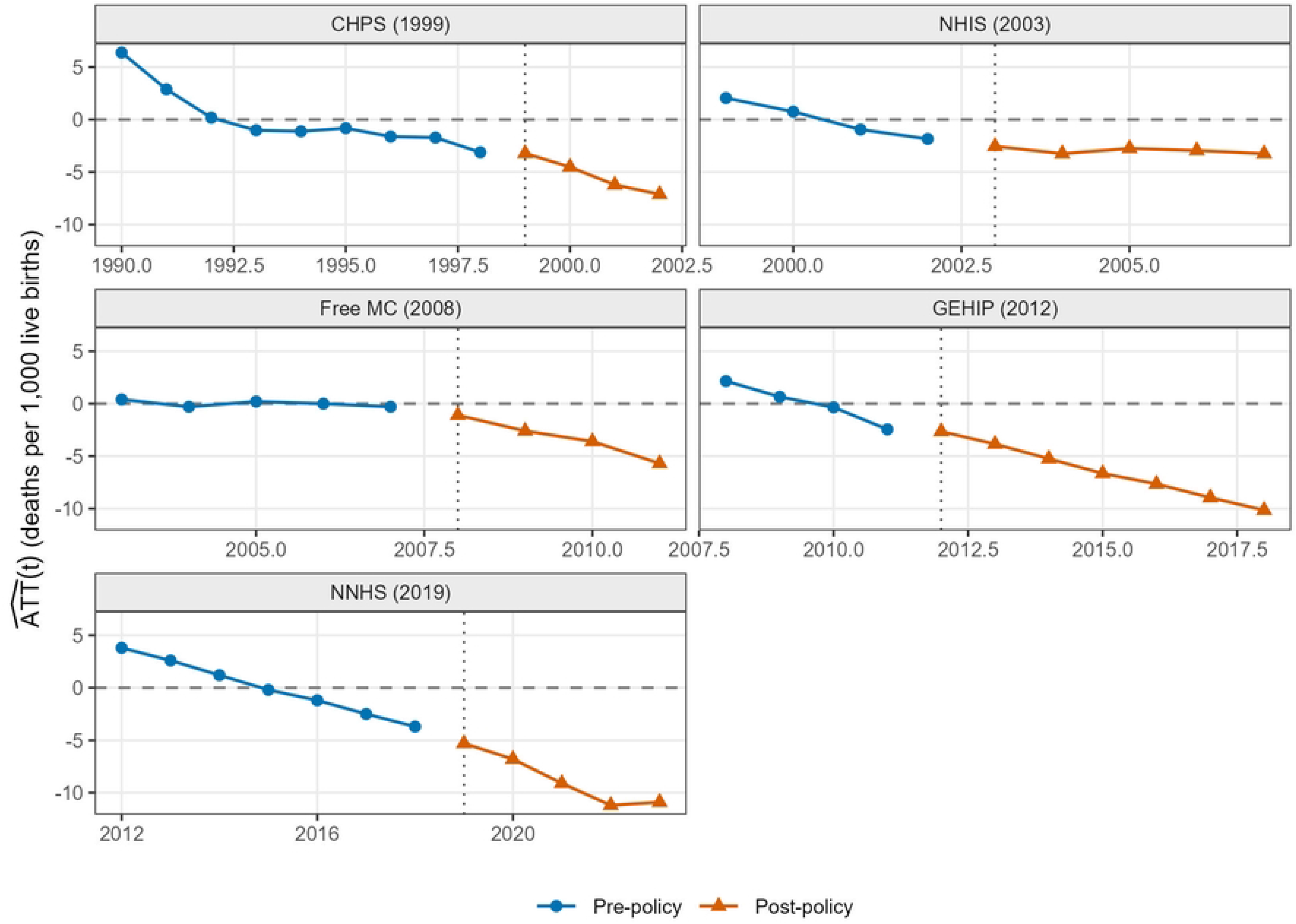
Event-Study DiD Estimates - Child Mortality Rate vs. Global Average

**Figure 4:**
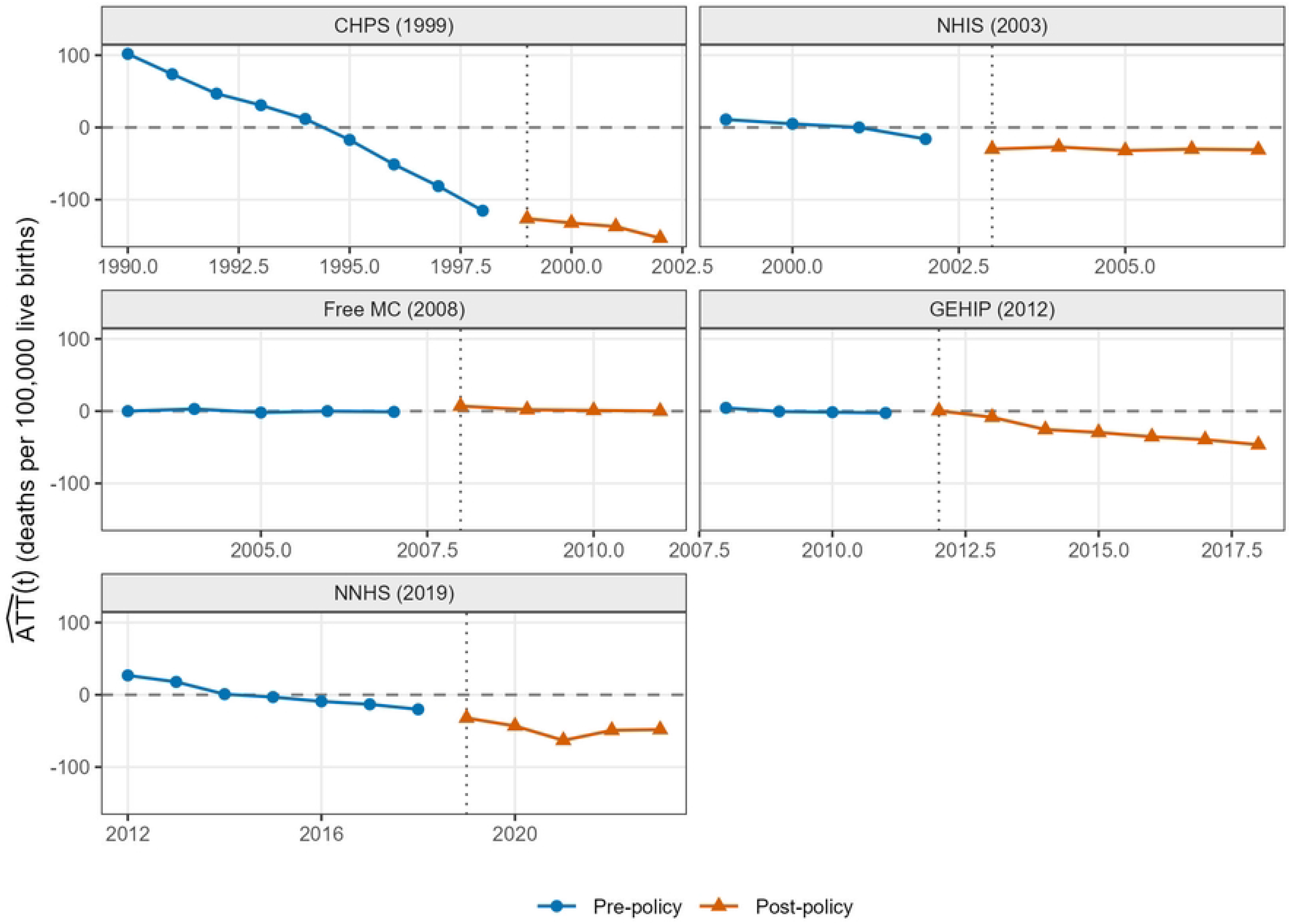
Event-Study DiD Estimates - Maternal Mortality Ratio vs. Global Average

Figure 3 displays the year-by-year ATT for child mortality rate against the global average. Pre-policy series are close to flat for NHIS, Free MC, and GEHIP, supporting the parallel trends assumption for those windows. GEHIP shows the sharpest post-treatment shift, with the ATT falling from near zero to around ― 10 by 2018. NNHS accelerates after 2019 but a mild pre-existing downward drift means its estimate is an upper bound. The CHPS pre-period slopes continuously downward through 1998 and continues unchanged after 1999, indicating an ongoing trend rather than a policy-induced break.

Figure 4 shows more variation in pre-trend behavior for MMR. The CHPS panel is the clearest violation: the ATT falls from + 101.8 in 1990 to ― 115.2 by 1998 at a slope of roughly ― 27 per year, continuing unchanged through 1999. This confirms that the large CHPS MMR estimate captures a pre-existing decline, and not a policy effect. Free MC sits near zero throughout its entire window, with no shift after 2008, consistent with evidence that access reforms require adequate facility capacity to reduce mortality [25]. GEHIP shows the flattest pre-trend of any MMR window, oscillating near zero from 2008 to 2011 before stepping steadily downward from 2012 onward, making it the most credible MMR estimate. NNHS accelerates sharply after 2019 but partly reflects COVID-19 disruption, and the mild pre-existing drift makes it an upper bound.

## Discussion

Ghana’s experience between 1990 and 2023 documents a country that reduced child mortality by roughly 72 percent and maternal mortality by roughly 75 percent over three decades of sustained policy effort. This paper asked a sharper question than prior descriptive work: which of the five major health interventions introduced between 1999 and 2019 can be credited with accelerating those declines, and by how much?

The staggered 2×2 Difference-in-Difference estimates, assessed against both the global average and the lower-middle-income country group, point to a consistent pattern. For child mortality, the Ghana Essential Health Interventions Program (GEHIP, 2012) and the National Newborn Health Strategy (NNHS, 2019) produce the largest and most consistent negative ATTs across both primary comparators. GEHIP is estimated to have reduced child mortality rate by approximately 6.5 deaths per 1,000 live births relative to the global trend, equivalent to 9.5 percent of Ghana’s pre-GEHIP mean, and by 3.2 deaths relative to the lower-middle-income group. NNHS shows a larger percentage effect at 16.4 percent against the global comparator, but the event-study path reveals a mild pre-existing downward drift in that window, so the estimate is best read as an upper bound on the program’s own contribution. The earlier policies produce more modest or ambiguous child mortality rate effects: Free MC (2008) at 4.0 percent relative to global and 1.5 percent relative to the income group, NHIS (2003) at 3.0 percent against global but near zero against the income group, and CHPS (1999) showing no Ghana-specific gain relative to lower-middle-income peers.

For maternal mortality, the GEHIP window again provides the most credible identification. The pre-policy ATT series for that window is nearly flat from 2008 to 2011 before stepping downward steadily from 2012 onward, and the estimated reduction of 7.2 percent relative to the global average and 5.2 percent relative to the lower-middle-income group is defensible given this pre-trend behavior. The CHPS (1999) estimate for maternal mortality is large in magnitude at 21.0 percent, but the event-study plot shows an uninterrupted downward slope from 1990 through 2002 that does not change at the 1999 policy marker, indicating the estimate captures a pre-existing trend rather than a policy effect. The Free Maternal Care policy (2008) produced no detectable change in Ghana’s maternal mortality trajectory relative to either comparator, with the annual ATT hovering near zero throughout the window. This finding aligns up with Srofenyoh et al. [10], who documented that emergency obstetric care capacity at the facility level remained a binding constraint through this period. Removing financial barriers to attendance is not sufficient when the facilities women attend lack the trained staff, blood supplies, and surgical capacity to manage obstetric emergencies.

Three broader lessons follow from these estimates. First, supply-side and quality-focused interventions appear to have driven Ghana’s most credible mortality improvements. GEHIP’s bundled approach to facility strengthening and service integration at the district level produced measurable effects on both child mortality rate and MMR, and the EmONC quality improvement documented at Ridge Regional Hospital illustrates the same logic: process changes within facilities translate into reduced case fatality rates in ways that financial access reforms alone do not [10]. Second, parallel trends behavior differs substantially across windows and outcomes, and any single-number summary of policy effects obscures this heterogeneity. The CHPS and NNHS windows for MMR both carry pre-trend violations that require the estimates to be interpreted as bounds rather than point estimates of causal effects. A companion analysis using disaggregated district-level data, where available, would allow formal pre-trend testing and tighter inference. Third, the sequential rollout of policies means that later estimates are identified against a background in which earlier programs are already operating. The GEHIP and NNHS effects therefore represent the marginal contribution of those programs above and beyond whatever baseline the NHIS, Free MC, and other programs had already established.

Ghana continues to fall short of SDG 3 maternal and child mortality targets on current trends. The evidence in this paper suggests that the most productive direction for further investment is on the supply side: strengthening facility capacity to manage obstetric emergencies, scaling the district-level service integration model that GEHIP piloted in the Upper West Region, and expanding the clinical quality protocols associated with the National Newborn Health Strategy. Financial access reforms remain necessary to ensure women reach facilities in the first place, but this analysis adds to the body of evidence that access without quality delivers limited mortality gains [25].

## Data Availability

All data used in this study are publicly available. Under-five mortality and maternal mortality estimates were obtained from the World Bank World Development Indicators database (https://databank.worldbank.org/source/world-development-indicators), drawing on modeled series produced by the UN Inter-agency Group for Child Mortality Estimation (https://childmortality.org) and the Maternal Mortality Estimation Inter-Agency Group (https://www.who.int/teams/maternal-newborn-child-adolescent-ageing/epidemiology-and-demography/maternal-mortality). No primary data were collected.

## Reflexivity Statement

Both first and second authors are Ghanaian nationals who completed their undergraduate education in Ghana before pursuing postgraduate training at US institutions. Our firsthand familiarity with Ghana’s health system and policy environment informs the contextual grounding of this research. Our training in applied econometrics shapes our methodological approach, favouring quantitative causal identification. We believe our dual positioning as Ghanaians with insider knowledge of the country’s health policy landscape and as researchers trained in rigorous causal inference strengthens both the contextual and analytical credibility of this study.

## Ethical Approval

Ethical approval for this type of study is not required by our institute.

## Funding

This research received no specific grant from any funding agency in the public, commercial, or not-for-profit sectors.

## Data Availability

No data was generated by this study. The following existing data sources were used: Mortality rate, under-5 (per 1,000 live births) [indicator SH.DYN.MORT] and Maternal mortality ratio, modeled estimate (per 100,000 live births) [indicator SH.STA.MMRT] from the World Bank World Development Indicators database, available via https://data.worldbank.org/indicator/SH.DYN.MORT and https://data.worldbank.org/indicator/SH.STA.MMRT.

https://data.worldbank.org/indicator/SH.DYN.MORT

## Acknowledgements

We thank Anthony Gariba for his advice and guidance in the development of this work. **Use of AI:** The authors used Claude (Anthropic) to assist with grammatical checking and modification of code. All AI-generated suggestions were reviewed and verified by the authors. The authors take full responsibility for the accuracy and integrity of all content in this manuscript.

## Conflicts of interest

The authors declare no conflict of interest.

## Appendix

**Table A1: Robustness — Sub-Saharan Africa Comparator**

**Table A1:**
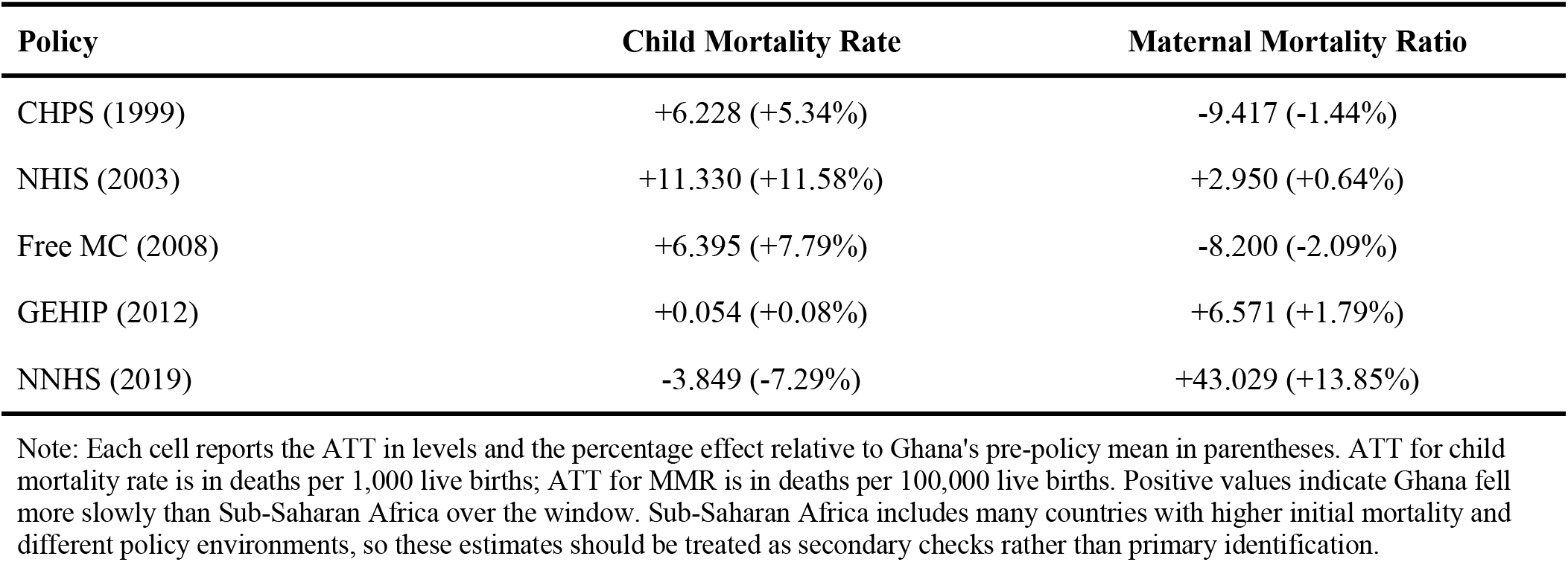
Robustness: DiD Estimates Using Sub-Saharan Africa (SSF) as Comparator.

| Policy | Child Mortality Rate | Maternal Mortality Ratio |
| --- | --- | --- |
| CHPS (1999) | +6.228 (+5.34%) | -9.417 (-1.44%) |
| NHIS (2003) | +11.330 (+11.58%) | +2.950 (+0.64%) |
| Free MC (2008) | +6.395 (+7.79%) | -8.200 (-2.09%) |
| GEHIP (2012) | +0.054 (+0.08%) | +6.571 (+1.79%) |
| NNHS (2019) | -3.849 (-7.29%) | +43.029 (+13.85%) |

Note: Each cell reports the ATT in levels and the percentage effect relative to Ghana's pre-policy mean in parentheses. ATT for child mortality rate is in deaths per 1,000 live births; ATT for MMR is in deaths per 100,000 live births. Positive values indicate Ghana fell more slowly than Sub-Saharan Africa over the window. Sub-Saharan Africa includes many countries with higher initial mortality and different policy environments, so these estimates should be treated as secondary checks rather than primary identification.

The SSF results for child mortality rate show positive ATTs for CHPS, NHIS, Free MC, and GEHIP, meaning Ghana fell more slowly than Sub-Saharan Africa for all but the NNHS window. This pattern reflects the fact that many Sub-Saharan African countries significantly reduced their child mortality rates during the 2000s under global child health programs including the Millennium Development Goals campaign, so the SSF group is not a stable counterfactual for Ghana over this period. The SSF-based child mortality rate estimates show a similar pattern: Ghana underperforms relative to the regional group for NHIS, GEHIP, and NNHS, which is consistent with the broader regional convergence documented in the descriptive statistics. The SSF comparator therefore provides limited identification value for this study, and the main conclusions should rest on the WLD and LMC estimates in Table 4.

---

**Event-Study Paths Against Lower Middle Income Comparator**

**Figure A1:**
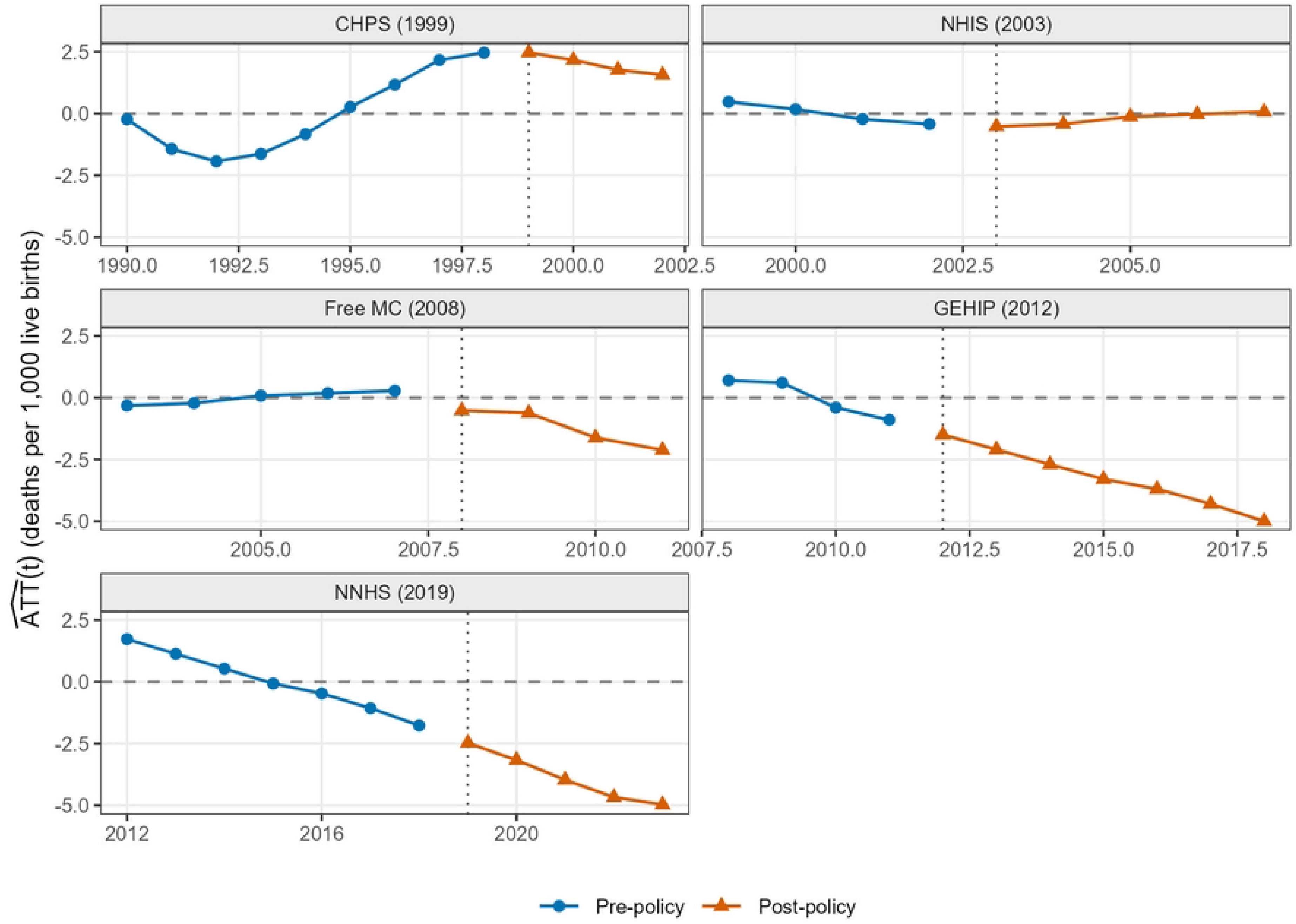
Event-Study Annual DiD Estimates: Child Mortality Rate vs. Lower Middle Income Average, by Policy Window\ Note: Annual ATT computed against the Lower Middle Income (LMC) comparator. Five-panel chart with one panel per policy window. Blue circles = pre-policy years; orange triangles = post-policy years. The dashed horizontal line marks zero; the dotted vertical line marks the policy year. Data source: World Bank WDI (SH.DYN.MORT).

**Figure A2:**
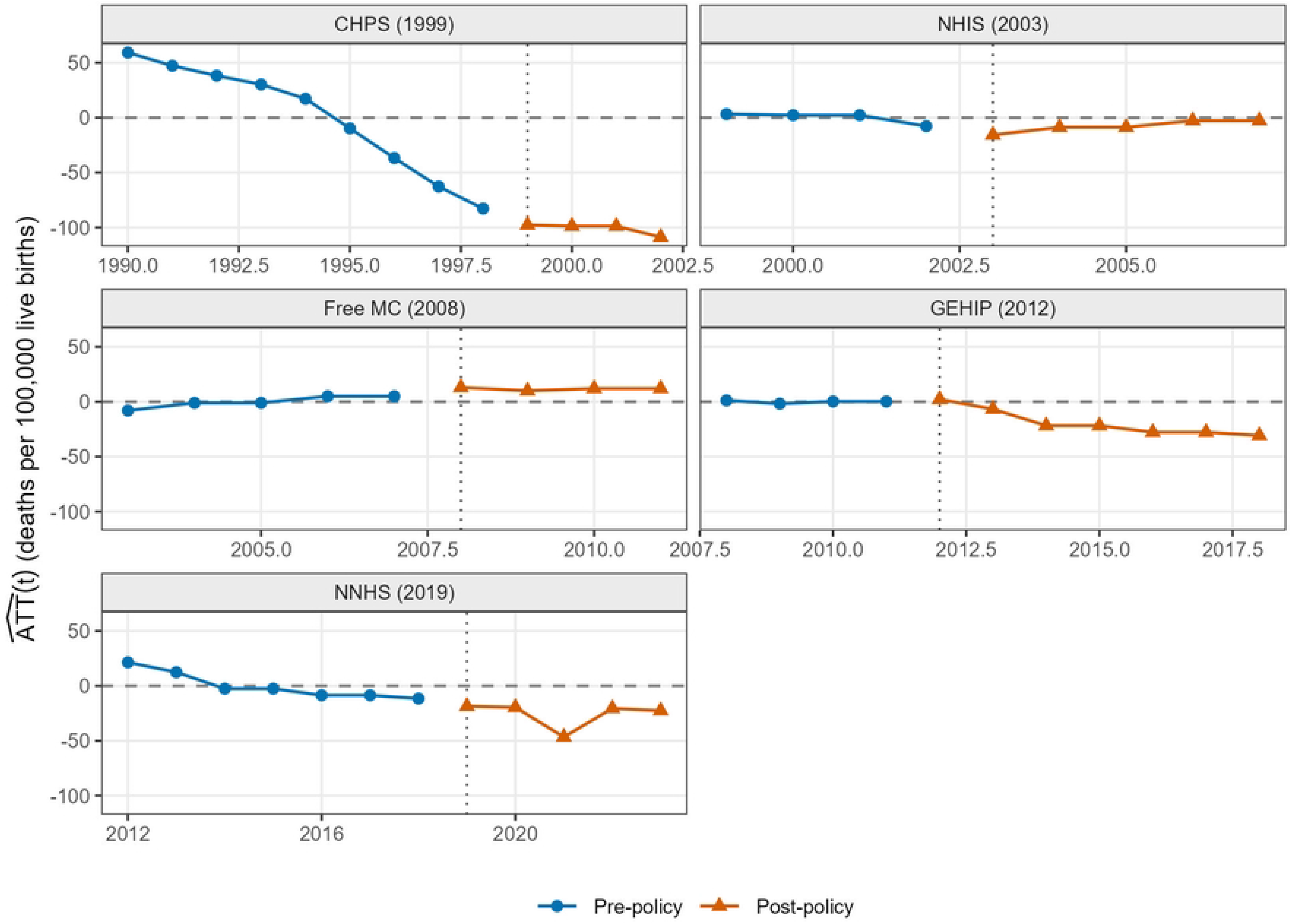
Event-Study Annual DiD Estimates: Maternal Mortality Ratio vs. Lower Middle Income Average, by Policy Window *Note: Annual ATT computed against the Lower Middle Income (LMC) comparator. Five-panel chart with one panel per policy window. Blue circles = pre-policy years; orange triangles = post-policy years. The dashed horizontal line marks zero; the dotted vertical line marks the policy year. Data source: World Bank WDI (SH.STA.MMRT)*.

## List of Abbreviations

Abbreviation: Full Term
ADF: Augmented Dickey-Fuller test
ANC: Antenatal Care
ARIMA: AutoRegressive Integrated Moving Average
ATT: Average Treatment Effect on the Treated
CHPS: Community-Based Health Planning and Services
COVID-19: Coronavirus Disease 2019
DHS: Demographic and Health Survey
DiD: Difference-in-Difference
EmONC: Emergency Obstetric and Newborn Care
FANC: Focused Antenatal Care
Free MC: Free Maternal Care Policy
GDP: Gross Domestic Product
GEHIP: Ghana Essential Health Interventions Program
HIV: Human Immunodeficiency Virus
IGME: UN Inter-agency Group for Child Mortality Estimation
JB: Jarque-Bera normality test
KPSS: Kwiatkowski-Phillips-Schmidt-Shin test
LMC: Lower Middle Income Countries (World Bank income group; primary comparator)
MDG: Millennium Development Goals
MMR: Maternal Mortality Ratio
NAS: National Ambulance Service
NHIS: National Health Insurance Scheme
NNHS: National Newborn Health Strategy
PP: Phillips-Perron test
PPH: Postpartum Haemorrhage
SDG: Sustainable Development Goals
SSF: Sub-Saharan Africa (World Bank country group code; robustness comparator)
TWFE: Two-Way Fixed Effects
U5MR: Under-Five Mortality Rate
UFEDC: User Fees Exemption for Delivery and Care
UN: United Nations
UNFPA: United Nations Population Fund
UNICEF: United Nations Children’s Fund
WDI: World Development Indicators (World Bank database)
WHO: World Health Organization
WLD: World (Global Average; World Bank country group code; primary comparator)

## Notes

### Competing Interest Statement

The authors have declared no competing interest.

### Author Declarations

This study used only publicly available, aggregated secondary data from the World Bank World Development Indicators database. No human participants were recruited and no individual-level or identifiable data were used. Therefore, IRB approval was not required.

